# Predictors of quality of life among Brazilian medical students during second coronavirus wave

**DOI:** 10.1101/2021.10.27.21265581

**Authors:** Filipe Silva Barreto, Ankilma do Nascimento Andrade Feitosa, Raquel Prudente de Carvalho Baldaçara, Leonardo Baldaçara, Elisangela Vilar de Assis, José Bruno Nunes Ferreira Silva

**Affiliations:** Curso de Medicina, Universidade Federal do Tocantins, 77001-923 Palmas, TO, Brasil; Curso de Medicina, Faculdade Santa Maria, 58900-000 Cajazeiras, PB, Brasil; Curso de Medicina, Universidade Federal de Campina Grande, 58900-000 Cajazeiras, PB, Brasil

**Author notes:** Corresponding author: José Bruno Nunes Ferreira Silva; address: Universidade Federal do Tocantins – Campus Universitário de Palmas – Avenida NS 15, ALCNO 14, Bala I, 109 Norte – Plano Diretor Norte - CEP: 77001-090 – Palmas (TO), Brasil.

**Keywords:** Mental health, COVID-19 pandemic, universities, women, sexual minorities, mobile applications

## Abstract

**Objective:** To assess the impact of COVID-19 second wave on quality of life (QoL) of Brazilian medical students.

**Methods:** The data from 210 medical students were collected through a web-based survey between February and March 2021. We assessed information about sociodemographic characteristics, health conditions, electronics tools-related to physical activity (PA-eTool), and health-associated QoL. Multiple linear regression was performed to identify the potential QoL predictors.

**Results:** The psychological domain was the most affected, showing lower score during COVID-19 second wave. Being male was a positive predictor in the physical health domain. Spending too much time indoors and sexual minority-group membership were associated with lower perception on psychological well-being. Personal benefit from having PA-eTool accessed and higher income were positively associated with psychological and environmental domains, respectively, and in the overall QoL.

**Conclusions:** Psychological well-being impairment is a long-term negative impact among Brazilian medical students during COVID-19. By evidencing that students’ QoL has been worsened in this global crisis, supportive interventions provided by community and universities are required to aid those vulnerable students facing mental health or QoL impairment.

## Introduction

For Brazil, the novel coronavirus disease or COVID-19 has posed a significant public health challenge ever since its emergence—and it seems to be far from over. As of October 8, 2021, a total of 21,532,558 cases and 599,810 fatalities have been reported in the country.^1^ In response to the crisis, the Brazilian Ministry of Health has released guidelines for mitigating the spread of the severe acute respiratory syndrome coronavirus 2 (SARS-CoV-2), which include avoiding crowds, wearing masks, maintaining physical distance from others, and taking the COVID-19 vaccine.^2^

Even as governments have made continual adjustments to protect people and societies from the coronavirus, studies have assessed its immediate and long-term effects on people’s mental health and well-being, which have a bearing on their day-to-day lives. One study has found people at risk of depression, anxiety, emotional disturbance, and physical impairment due to a compromised quality of life (QoL) amid the pandemic.^3^ University students have been shown to be particularly vulnerable to the negative impacts of COVID-19.^4^ During the first wave of COVID-19, we reported on the issues faced by medical students during social isolation which influenced their healthy habits and QoL.^5^

When a second wave of infections swept Brazil at the end of 2020, the number of cases rose among younger ages.^6^ Thus, we became interested in expanding our assessment to gain a clearer picture of the long-term effects of COVID-19 on medical students’ QoL. So far, little is known about the long-term psychological impact of COVID-19 on Brazilian medical students and coping strategies for maintaining a positive QoL amid the global health crisis. Therefore, we decided to continue assessing medical students’ QoL to understand the long-term effects of COVID-19 with a view to guide future wellness programs.

## Methods

### Study Design

Data was collected for this cross-sectional study from February to March 2021, the period that corresponded with the second wave of COVID-19.^7^ All medical students from the Federal University of Tocantins in Palmas, Federal University of Campina Grande, and Santa Maria College, both in Cajazeiras were eligible to participate. A self-reported e-questionnaire (Google Forms^®^) was promoted with links to the form sent by email, WhatsApp, Facebook, and Instagram. To avoid multiple responses submitted by the same subject, participants were asked to include their email addresses. Participation was voluntary and completion of the survey implied consent. Respondents were excluded if they were not undergraduate medical students at universities included in the study or if they did not complete the questions properly. Of the 233 men and women aged 18 years or over who answered the e-questionnaire, 23 (9.87%) were excluded. The final sample (n=210) comprised 20.8% of the students enrolled in those courses. The National Research Ethics Commission of the Ministry of Health (CONEP) (C.A.A.E.:33567120.2.1001.5519) approved the study.

The participants provided sociodemographic data (age, sex, sexual orientation, marital status, smoking habits, and alcohol intake), respiratory disease history, and details on PA-eTool access. They also provided information about their body weight and height, which are used to calculate body mass index (BMI), along with the amount of time they spent indoors. Details of design and characteristics of the study can be found in our previous publication.^5^ In order to assess the QoL, we used the World Health Organization’s Quality of Life (WHOQOL-Bref) questionnaire and generated four domain scores: physical health, psychological well-being, social relationships, and environment.^8^

### Data Analysis

The explanatory variables related to the sociodemographic profile, PA-eTool use, and respiratory diseases were expressed as relative and absolute frequencies. WHOQOL-Bref scores were obtained by calculating the mean value of the answers for each of the four areas. The overall QoL and individual domain scores ranged from 0 to 100. Independent Student’s t-tests and one-way analysis of variance (ANOVA) were used to examine differences in each variable’s scores (Table S1 available as online supplementary material). The Cronbach’s alpha value for the WHOQOL-Bref questionnaire used in this study was 0.891, which was deemed acceptable. The internal consistency coefficient was not significantly reduced when the items in each domain were excluded. Spearman’s rank correlation and Friedman’s/Dunn’s tests were performed to determine the level of agreement between the domains. At the end, exploratory variables (p<0.2 in the univariate analysis) were inserted into the multiple linear regression models and performed for each WHOQOL-Bref domain. The data was analyzed with a significance level of p ≤ 0.05 using SPSS Statistics software version 25 (IBM Corp., Armonk, NY, USA).

## Results

Table S1 shows the sociodemographic characteristics of the participants. Most of the medical students were young adults, female, heterosexual, and single. Most had received more than 3 salaries at least, were non-smokers, non-alcohol users, and did not have respiratory disease. Of the participants, 74.28% reported spending more than 16 hours indoors during the second COVID-19 wave. Weight changes were reported by 71.9%, with most of them declaring they had gained weight. As shown, respondents who were obese and overweight accounted for just over 30% of the sample and 40.5% reported using the PA-eTool (Table S1).

As shown in Table 1, the scores of each WHOQOL-Bref domain showed a statistically significant correlation between the domains and overall QoL scores. The strongest correlation was found in the psychological well-being domain (p<0.001, r = 0.831), which had the lowest mean score (60.09 ± 16.00) during the second COVID-19 wave. The overall QoL scores of medical students were 64.67 ± 13.14, with a median of 65.75 points and scores ranging from 26.50 to 97.

**Table 1.** Descriptive and inferential analysis of the WHOQOL-Bref (World Health Organization’s Quality of Life) among Brazilian medical students facing second wave of COVID-19.

| | Mean ± SD | Correlation with WHOQOL-Bref |
| --- | --- | --- |
| WHOQOL-Bref | 64.67 ± 13.14 * | |
| D1 physical health | 67.32 ± 15.06 * | p < 0.001 (r = 0.763) ** |
| D2 psychological well-being | 60.09 ± 16.00 * | p < 0.001 (r = 0.831) ** |
| D3 social relationships | 62.20 ± 21.04 * | p < 0.001 (r = 0.770) ** |
| D4 environmental | 69.06 ± 15.13 * | p < 0.001 (r = 0.694) ** |
|  | p < 0.001 |  |

\*Friedman's/Dunn's test (D1 vs. D2, p < 0.001; D1 vs. D3, p = 0.001; D1 vs. D4, p = 1.000; D2 vs. D3, p = 0.513; D2 vs. D4, p < 0.001; D3 vs. D4, p < 0.001).
\*\*Spearman's rank correlation with the overall domain (D).

According to the univariate analysis shown in Table S1, heterosexuals and non-smokers had a higher mean for the physical and psychological well-being domains (p<0.05). Women and sexual minority group membership was associated with lower scores in the physical and environmental domains, respectively (p<0.05). For the social domain, being obese or underweight translated into a lower mean (p<0.05). Those who earned over three Brazilian minimum wages had higher scores in the environmental domain (p<0.001). Assessing workout/fitness applications (apps) for PA was positively associated with a higher mean for the psychological domain and overall QoL. Similarly, overall QoL values were higher for heterosexuals and those with up to three minimum wages (p<0.01), differing only in the BMI index, with underweight and obese students having a lower mean (p<0.05).

The results of the multiple linear regression analysis are presented in Table 2. The gender variable only remained associated with the physical domain, with women showing a lower perception of QoL than men. Spending more than 16 hours indoors and sexual minority group membership were negatively associated with the psychological domain. No predictors were found in the social domain. Family income for those who had earned a minimum of three wages was shown to be a positive predictor of the environment domain of QoL. Use of apps, such as the PA-eTool, was also a positive predictor of the psychological domain of QoL as well as the overall QoL.

**Table 2.**
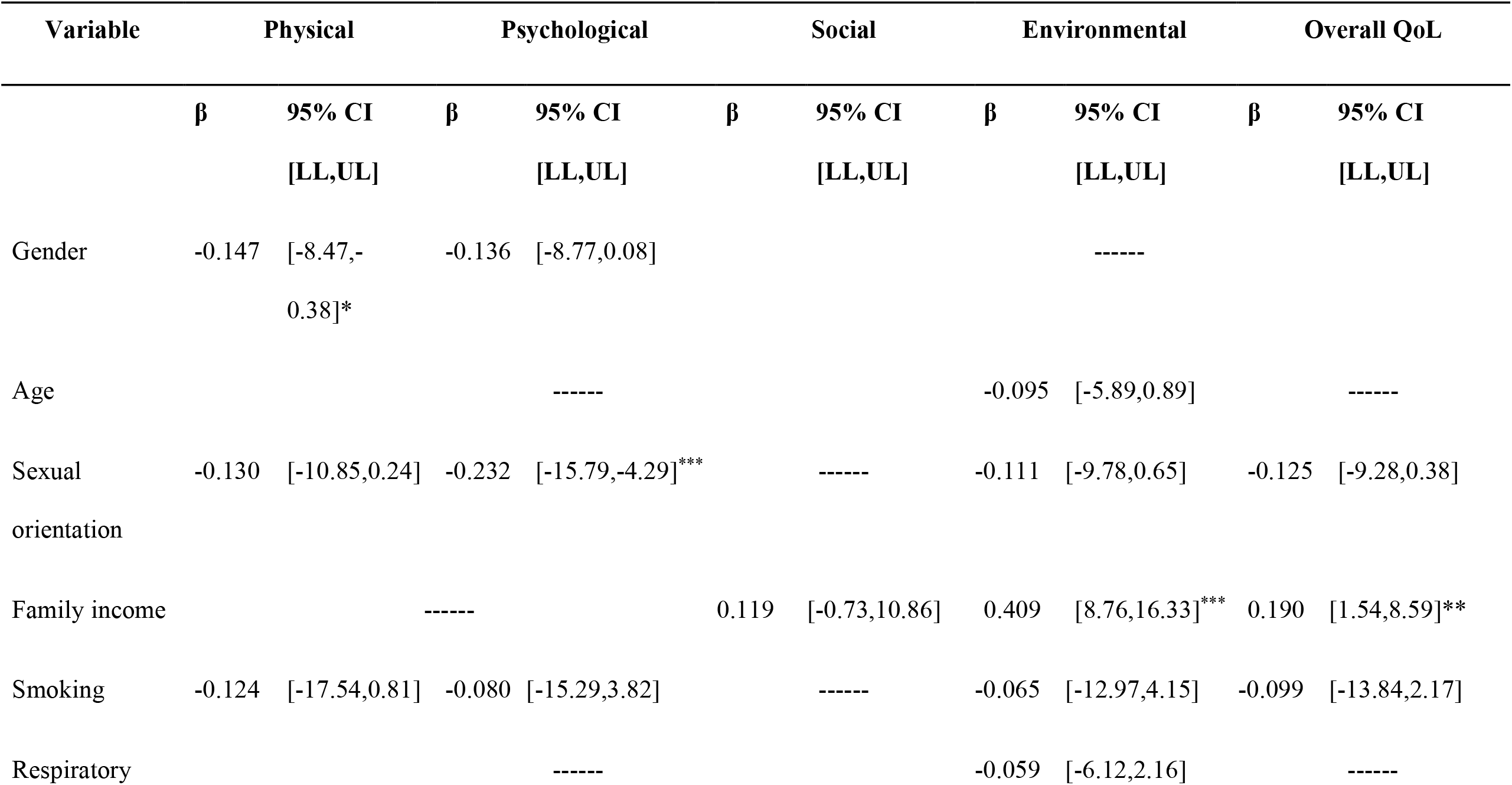
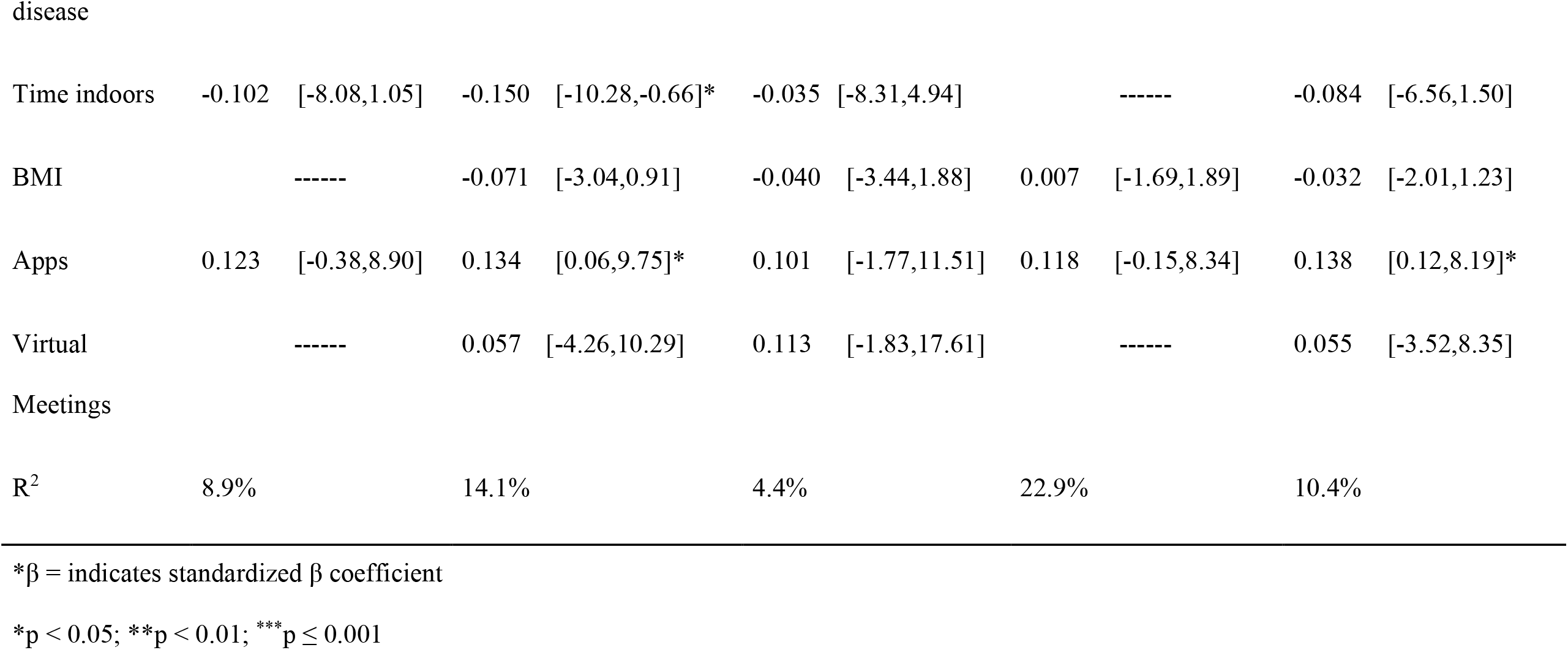
Multiple Linear Regression Analysis for WHOQOL-Bref (World Health Organization’s Quality of Life) domains during the second wave of COVID-19 in Brazil.

| Variable | Physical |  | Psychological |  | Social |  | Environmental |  | Overall QoL |  |
| --- | --- | --- | --- | --- | --- | --- | --- | --- | --- | --- |
| | $\beta$ | 95% CI<br>[LL,UL] | $\beta$ | 95% CI<br>[LL,UL] | $\beta$ | 95% CI<br>[LL,UL] | $\beta$ | 95% CI<br>[LL,UL] | $\beta$ | 95% CI<br>[LL,UL] |
| Gender | -0.147 | [-8.47,-0.38]* | -0.136 | [-8.77,0.08] |  |  |  | ----- |  |  |
| Age |  |  |  | ----- |  |  | -0.095 | [-5.89,0.89] |  | ----- |
| Sexual orientation | -0.130 | [-10.85,0.24] | -0.232 | [-15.79,-4.29]*** |  | ----- | -0.111 | [-9.78,0.65] | -0.125 | [-9.28,0.38] |
| Family income |  |  | ----- |  | 0.119 | [-0.73,10.86] | 0.409 | [8.76,16.33]*** | 0.190 | [1.54,8.59]** |
| Smoking | -0.124 | [-17.54,0.81] | -0.080 | [-15.29,3.82] |  | ----- | -0.065 | [-12.97,4.15] | -0.099 | [-13.84,2.17] |
| Respiratory |  |  |  | ----- |  |  | -0.059 | [-6.12,2.16] |  | ----- |
| Time indoors | -0.102 | [-8.08,1.05] | -0.150 | [-10.28,-0.66]* | -0.035 | [-8.31,4.94] |  | ----- | -0.084 | [-6.56,1.50] |
| BMI |  | ----- | -0.071 | [-3.04,0.91] | -0.040 | [-3.44,1.88] | 0.007 | [-1.69,1.89] | -0.032 | [-2.01,1.23] |
| Apps | 0.123 | [-0.38,8.90] | 0.134 | [0.06,9.75]* | 0.101 | [-1.77,11.51] | 0.118 | [-0.15,8.34] | 0.138 | [0.12,8.19]* |
| Virtual Meetings |  | ----- | 0.057 | [-4.26,10.29] | 0.113 | [-1.83,17.61] |  | ----- | 0.055 | [-3.52,8.35] |
| R<sup>2</sup> | 8.9% |  | 14.1% |  | 4.4% |  | 22.9% |  | 10.4% |  |

\*β = indicates standardized β coefficient
\*p < 0.05; \*\*p < 0.01; \*\*\*p ≤ 0.001

## Discussion

This study found that the mental health of medical students was affected during the second COVID-19 wave, particularly those who spent more than 16 hours indoors and identified themselves as gay, lesbian, or bisexual. During the first coronavirus wave, our preliminary survey had demonstrated that students who spent less than 12 hours indoors had a positive perception in the psychological domain.^5^ Lately, the relaxation of social distancing policies has allowed medical students to return to their routine activities. In addition, access to accurate knowledge of COVID-19 has reduced their sense of uncertainty and fear. However, individuals facing social isolation are still experiencing negative changes in their well-being because they lack freedom, are far from their loved ones, and also, are frequently bored at home.^9^ Apart from constantly facing social exclusion, sexual minority medical students may have to deal with rejection and discrimination upon returning to the negative family nucleus due to the closure of higher education institutions. Moreover, Brazilian young adults have reported feeling “suffocated” as they are being forced to hide their truth by not expressing their sexual identity in the same environment as potential aggressors since it may result in psychological damage.^10,11^

Consistent with our findings, a meta-analysis performed by Solis and Lotufo-Neto^12^ has found that being female and having low socioeconomic status are predictors of negative QoL among Brazilian medical students. In addition, a study by Hung et al.^13^ provided evidence that impairment of physical health in women is a long-term consequence of stress, anxiety, and depression experienced during the COVID-19 pandemic. When women experience domestic violence and economic or educational disadvantage, they are more vulnerable to mental disorders.^14^ Additionally, becoming a mother has been associated with higher likelihood of depression and anxiety during the second wave of COVID-19 in Brazil.^15^ Furthermore, it has been found that higher frequency of mood disorders in women has been associated with lower QoL^12^; however, gender equality and women’s empowerment may be effective strategies to support them during the pandemic.^16^

Psychologically speaking, losing a job increases stress and mental health disorders in people.^16^ In Brazil, millions have become unemployed because of the disruption of economic activities due to COVID-19. Even though an emergency support income was provided by the Brazilian government during the pandemic waves, a low-income household environment may affect the QoL, and feelings of poverty can worsen mental health.

In the age of advanced technology, accessing the Internet has become a day-to-day reality for supporting remote work and education. In this study, we also considered digital tools as a coping strategy for QoL. As evidenced in the first wave^5^, using workout or fitness apps remained a positive predictor for protecting the psychological conditions and QoL of medical students. However, online home-based exercise monitored by physical education teachers was not associated with QoL in the current wave. One possible reason was that students could practice physical activities at fitness and gym centers due to the relaxation of protective measures. Our data emphasizes the importance of creating mechanisms to provide regular PA programs to protect QoL during the pandemic.

Practicing PA, maintaining an improved lifestyle through healthy eating habits, reducing alcohol intake, and better sleep quality along with employing complementary and alternative interventions have been suggested as methods to prevent psychological issues following traumatic periods during and after COVID-19 pandemic.^17^ Kleemann et al.^18^ emphasized that PA practice can be used as a complementary therapeutic approach and be recommended to patients with mental illness. Additionally, telemedicine can be a strategy for supporting face-to-face patient encounters in the long-term by facilitating dissemination of general mental health recommendations.^5,17^

Addressing the possible limitations of this study, data collected from a specific group of undergraduate students may not be representative of all Brazilian medical students. Our data suggested findings similar to those of a study in which Brazilian female students were more likely to experience negative psychological issues during COVID-19 pandemic.^19^ Our convenience sampling strategy could have affected the size sample, limiting the participation of students who do not have internet access. In addition, a cross-sectional study offers limited ability to make causal inferences, thus we intend to continue investigating these issues raised due to the long-term consequences of the ongoing COVID-19 pandemic.

This study highlighted the long-term impact of COVID-19 pandemic on the quality of life of Brazilian medical students. The findings from this study provide evidence that the second wave affected the environmental, physical, and psychological health of medical students. However, the PA-eTool was an element of higher self-perception of well-being. Our data present elements for calling the universities to adopt supportive actions to protect vulnerable students from facing mental health or quality of life impairment during critical events, such as pandemics.

## Data Availability

All data produced in the present study are available upon reasonable request to the authors

## Acknowledgments

The authors would like to thank UFT for the support given to the development of this research work. FSB received scholarship from CNPq (Brazilian National Council for Scientific and Technological Development) which is gratefully acknowledged.

