## Supplemental Table 1 for "Predictors of quality of life among Brazilian medical students during second coronavirus wave"

**Table S1. Effects of sociodemographic characteristics, health conditions, weight change, and eTools on the domains scores of the WHOQOL-Bref (World Health Organization's Quality of Life) among medical students facing second wave of COVID-19.**

| Variables | n (%) | Physical | Psychological | Social | Environmental | Overall QoL |
|---|---|---|---|---|---|---|
| Gender | | | | | | |
| Female | 114 (54.3) | 65.37 ± 14.79 | 58.59 ± 15.53 | 63.70 ± 19,33 | 70.12 ± 13.58 | 64.45 ± 12.05 |
| Male | 96 (45.7) | 69.65 ± 15.13 | 61.86 ± 16.44 | 60.41 ± 22.88 | 67.79 ± 16.76 | 64.93 ± 14.39 |
| | | p = 0.04 | p = 0.140 | p = 0.266 | p = 0.276 | p = 0.795 |
| Age | | | | | | |
| 18 - 19 | 16 (7.6) | 62.19 ± 17.30 | 55.88 ± 17.20 | 58.25 ± 24.95 | 69.25 ± 12.85 | 61.39 ± 15.52 |
| 20 - 24 | 131 (62.4) | 67.91 ± 14.17 | 60.63 ± 16.01 | 63.90± 20.28 | 70.76 ± 14.47 | 65.80 ± 12.43 |
| > 24 | 63 (30) | 67.41 ± 16.23 | 60.03 ± 15.76 | 59.65 ± 21.51 | 65.48 ± 16.53 | 63.14 ± 13.85 |
| | | p = 0.359 | p = 0.535 | p = 0.311 | p = 0.074 | p = 0.246 |
| Sexual orientation | | | | | | |
| Heterosexual | 176 (83.8) | 68.43 ± 15.00 | 61.93 ± 15.70 | 62.05 ± 21.20 | 69.99 ± 15.51 | 65.60 ± 13.53 |
| Sexual minority | 34 (16.2) | 61.62 ± 14.25 | 50.56 ± 14.20 | 62.97 ± 20.46 | 64.24 ± 12.06 | 59.85 ± 9.71 |
| | | p = 0.015 | p < 0.001 | p = 0.815 | p = 0.042 | p = 0.005 |
| Single | | | | | | |
| No | 20 (9.5) | 69.50 ± 14.46 | 64.40 ± 16.34 | 61.60 ± 21.40 | 67.00 ± 19.13 | 65.63 ± 14.70 |
| Yes | 190 (90.5) | 67.09 ± 15.14 | 59.63 ± 15.94 | 62.26 ± 21.06 | 69.27 ± 14.69 | 64.56 ± 13.01 |
| | | p = 0.498 | p = 0.206 | p = 0.895 | p = 0.524 | p = 0.732 |
| Family income | | | | | | |

| | | | | | | |
|---|---|---|---|---|---|---|
| Up to 3 | 87 (41.4) | 66.72 ± 16.06 | 59.83 ± 17.46 | 59.44 ± 22.22 | 61.38 ± 13.47 | 61.84 ± 14.01 |
| > 3 | 123 (58.6) | 67.75 ± 14.37 | 60.27 ± 14.95 | 64.15 ± 20.02 | 74.49 ± 13.87 | 66.66 ± 12.16 |
| | | p = 0.629 | p = 0.845 | p = 0.110 | p < 0.001 | p = 0.009 |
| Smoking habit | | | | | | |
| No | 199 (94.8) | 67.89 ± 14.91 | 60.64 ± 16.01 | 62.34 ± 21.22 | 69.38 ± 15.25 | 65.06 ± 13.22 |
| Yes | 11 (5.2) | 57.09 ± 14.68 | 50.09 ± 12.65 | 59.64 ± 18.11 | 63.18 ± 11.61 | 57.50 ± 9.41 |
| | | p = 0.020 | p = 0.033 | p = 0.680 | p = 0.186 | p = 0.063 |
| Alcohol consumption | | | | | | |
| No | 129 (61.4) | 67.49 ± 14.72 | 61.06 ± 15.69 | 62.72 ± 20.87 | 69.88 ± 15.14 | 65.29 ± 13.12 |
| Yes | 81 (38.6) | 67.06 ± 15.68 | 58.53 ± 16.46 | 61.36 ± 21.40 | 67.74 ± 15.09 | 63.67 ± 13.19 |
| | | p = 0.842 | p = 0.265 | p = 0.649 | p = 0.319 | p = 0.387 |
| Respiratory disease | | | | | | |
| No | 150 (71.4) | 67.95 ± 15.41 | 60.91 ± 16.36 | 62.43 ± 20.83 | 70.02 ± 15.08 | 65.33 ± 13.20 |
| Yes | 60 (28.6) | 65.75 ± 14.16 | 58.02 ± 14.99 | 61.62 ± 21.72 | 66.65 ± 15.09 | 63.01 ± 12.95 |
| | | p = 0.339 | p = 0.237 | p = 0.802 | p = 0.145 | p = 0.249 |
| Duration of time spent indoors | | | | | | |
| 8-12 h | 25 (11.9) | 67.92 ± 15.81 | 63.32 ± 17.17 | 58.32 ± 20.42 | 64.16 ± 18.47 | 63.43 ± 15.06 |
| 12-16 h | 29 (13.8) | 72.76 ± 13.35 | 66.52 ± 12.89 | 68.79 ± 15.74 | 69.24 ± 14.43 | 69.33 ± 11.53 |
| 16-20 h | 75 (35.7) | 67.00 ± 15.12 | 58.25 ± 15.39 | 58.73 ± 20.89 | 68.80 ± 15.03 | 63.20 ± 13.07 |
| 20-24 h | 81 (38.6) | 65.49 ± 15.15 | 58.48 ± 16.70 | 64.23 ± 22.48 | 70.74 ± 14.25 | 64.74 ± 12.98 |

| | | | | | | |
|---|---|---|---|---|---|---|
| | | p = 0.168 | p = 0.056 | p = 0.089 | p = 0.303 | p = 0.186 |
| Weight change | | | | | | |
| Increased | 103 (49) | 67.39 ± 14.46 | 60.24 ± 15.85 | 62.91 ± 20.75 | 68.80 ± 15.92 | 64.83 ± 13.32 |
| Stable | 59 (28.1) | 67.31 ± 16.04 | 62.12 ± 16.30 | 60.39 ± 21.54 | 68.95 ± 14.79 | 64.69 ± 13.09 |
| Decreased | 48 (22.9) | 67.21 ± 15.42 | 57.25 ± 15.84 | 62.88 ± 21.35 | 69.75 ± 14.03 | 64.27 ± 13.09 |
| | | p = 0.998 | p = 0.292 | p = 0.741 | p = 0.936 | p = 0.970 |
| BMI | | | | | | |
| Underweight | 6 (2.8) | 69.00 ± 11.30 | 62.33 ± 10.52 | 52.17 ± 18.39 | 64.67 ± 10.25 | 62.04 ± 7.10 |
| Normal | 140 (66.7) | 66.93 ± 15.34 | 60.11 ± 16.58 | 62.91 ± 20.64 | 69.04 ± 14.63 | 64.75 ± 13.18 |
| Overweight | 51 (24.3) | 69.65 ± 14.58 | 62.20 ± 14.92 | 64.88 ± 21.34 | 71.53 ± 16.12 | 67.06 ± 13.14 |
| Obesity | 13 (6.2) | 61.69 ± 15.06 | 50.54 ± 13.42 | 48.54 ± 21.16 | 61.62 ± 16.88 | 55.60 ± 11.81 |
| | | p = 0.364 | p = 0.130 | p = 0.048 | p = 0.171 | p = 0.042 |
| **Electronic tools for physical activity** | | | | | | |
| Youtube | | | | | | |
| Yes | 53 (25.2) | 68.96 ± 15.08 | 57.45 ± 16.66 | 62.28 ± 20.16 | 68.66 ± 13.53 | 64.34 ± 12.30 |
| No | 157 (74.8) | 66.77 ± 15.06 | 60.97 ± 15.72 | 62.17 ± 21.39 | 69.19 ± 15.67 | 64.78 ± 13.45 |
| | | p = 0.361 | p = 0.166 | p = 0.972 | p = 0.826 | p = 0.835 |
| Applications | | | | | | |
| Yes | 53 (25.2) | 70.15 ± 12.10 | 63.94 ± 15.15 | 66.79 ± 18.98 | 72.28 ± 13.85 | 68.29 ± 11.07 |
| No | 157 (74.8) | 66.37 ± 15.86 | 58.78 ± 16.11 | 60.64 ± 21.53 | 67.97 ± 15.42 | 63.44 ± 13.59 |
| | | p = 0.073 | p = 0.042 | p = 0.066 | p = 0.072 | p = 0.020 |

| | | | | | | |
|---|---|---|---|---|---|---|
| Virtual meetings | | | | | | |
| Yes | 21 (10) | 69.19 ± 15.27 | 64.67 ± 19.49 | 70.57 ± 20.35 | 70.29 ± 13.09 | 68.68 ± 13.78 |
| No | 189 (90) | 67.12 ± 15.06 | 59.58 ± 15.54 | 61.26 ± 20.96 | 68.92 ± 15.36 | 64.22 ± 13.03 |
| | | p = 0.551 | p = 0.167 | p = 0.054 | p = 0.696 | p = 0.141 |
